# Obesity Prevention Efforts in France, Italy, and Japan - A Scoping Review Protocol

**DOI:** 10.1101/2023.11.26.23299027

**Authors:** Nour M. Hammad, DeirDre K. Tobias

**Affiliations:** Department of Nutrition, Harvard T.H. Chan School of Public Health, Boston, MA, USA; Department of Medicine, Brigham and Women’s Hospital and Harvard Medical School, Boston, MA, USA

## Abstract

**Background:** Population-level prevention efforts in the US have yet to correct the upward trends in increasing US obesity prevalence. Italy, France, and Japan are high-income countries that have employed approaches aimed at obesity prevention that may have contributed to their favorable obesity trends. It is plausible that these efforts could be culturally adapted with similar effectiveness to the US population. This paper outlines the protocol of a scoping review that will identify and characterize the range of obesity prevention efforts implemented in Italy, France, and Japan to inform potential strategies for obesity prevention in the US.

**Methods and analysis:** We will conduct a systematic search in PubMed, Embase, and Cochrane. We will assess articles that describe any national, regional, or local public health and/or clinical guideline, recommendation, intervention, campaign, policy or other population-level effort targeting obesity prevention in Italy, France, and Japan. Two independent reviewers will screen the articles, and a third independent reviewer will resolve conflicts that arise upon screening. Descriptive synthesis will be performed to describe the country-specific prevention strategies and summarize the available evidence for their effectiveness.

**Discussion:** The purpose of this paper is to describe the study protocol for a scoping review that will provide a comprehensive summary of available evidence for obesity prevention strategies that have been implemented in Italy, France, and Japan. These strategies will inform potential prevention efforts for consideration in the US.

**Scoping review registration DOI:** https://doi.org/10.17605/OSF.IO/M35YX.

## Background

Obesity is a growing and multi-faceted public health problem in the US and globally, with prevalence reaching 41.9% of US adults.^1^ Obesity is associated with adverse physical health outcomes including mortality, cardiovascular diseases, cancer, and respiratory problems, underscoring the need for effective population-based strategies for prevention.^2^ Individuals with obesity are also at a higher risk of mental health conditions such as depression and anxiety, and are more likely to experience eating disorders, poorer self-esteem, and lower quality of life.^3,4^ Additionally, obesity places a huge burden on the US healthcare system. It is estimated that the US national cost of obesity in adults was $260.6 billion in 2016.^5^ Despite the awareness of the health, societal, and economic consequences of obesity, population-level prevention efforts have yet to correct the upward trends in increasing US obesity prevalence.

Italy, France, and Japan are high-income countries that have garnered attention for maintaining a low prevalence of obesity in recent decades, while neighboring countries have experienced increases.^6,7^ Notably, these countries employed approaches aimed at obesity prevention that may have contributed to their favorable obesity trends; it is plausible that these efforts could be culturally adapted with similar effectiveness to the US population.

## Methods

### Objectives

This scoping review aims to identify and characterize the range of obesity prevention efforts implemented in Italy, France, and Japan to inform potential strategies for obesity prevention in the US and elsewhere.

### Study designs

There will be no exclusions based on study design. All existing literature, including primary research studies, letters and editorials, systematic reviews, meta-analyses, Internet websites and blogs, national and/or local guidelines and policies, and presentations that describes and/or evaluates population-based interventions addressing obesity in Italy, France, and Japan will be assessed.

### Study populations

All general populations, with no age restrictions, living in the following settings (Italy, France, and Japan) and exposed to population-level obesity prevention efforts.

### Eligible interventions

Any national, regional, or local public health and/or clinical guideline, recommendation, intervention, campaign, policy or other population-level effort targeting obesity/weight prevention in Italy, France, and Japan.

### Outcomes of interest

Any indicators of obesity such as body mass index (BMI), body weight, waist circumference, at the individual level; population-level summary statistics including overweight and obesity prevalence, time trends, and changes over time (e.g., pre/post implementation of a policy).

### Electronic search strategies

We will conduct a systematic search in the following electronic databases, from inception to September 29, 2023: PubMed, Embase, and Cochrane. The search terms are detailed below the discussion section. The reference lists of accepted articles will be hand-screened for additional citations. Grey literature and websites of policies, public health campaigns, interventions, etc. that are unlikely to be published in peer reviewed journals will be searched via Google and Google Scholar.^8^

### Languages

There is no restriction on languages. Literature published in the English, Italian, French, and Japanese languages will all be considered for screening and inclusion.

### Inclusion and Exclusion Criteria

Studies will be included if they describe any population-level efforts targeting obesity/weight prevention in Italy, France, and Japan. Studies will be excluded if their outcomes were not related to obesity prevention, or the population was comprised exclusively of pregnant women or participants with comorbidities, or the obesity prevention efforts took place in settings exclusively not in Italy, France, and Japan, or the efforts had a short follow up duration of less than 6 months if applicable.

## Data collection and analysis

### Title and abstract screening phase

Initial screening of the individual titles and abstracts (where applicable) will be conducted by 2 independent reviewers using the Covidence systematic review software, Veritas Health Innovation, Melbourne, Australia. Available at www.covidence.org. A third independent reviewer will resolve conflicts as needed. Studies will be reviewed for eligibility and excluded if any exclusion criteria are met.

### Full text screening phase

Screening at this phase will be completed by 2 independent reviewers in Covidence. In this step, a reason for exclusion will be captured. All conflicts, including reasons for exclusion, will be resolved by a third independent reviewer as needed. The reference lists of accepted articles and relevant reviews will be screened to identify additional citations.

### Grey literature search

A grey literature search will be conducted alongside the published scientific literature screening. We will use Google and Google Scholar using an appropriate search term(s), and screening of the returned search results will be limited to the first 30 pages each.

## Data extraction and management

We will extract data for the final set of accepted articles (including grey literature) using a template developed and tested on Covidence. Specifically, we will extract:

1. Study characteristics
  a. Author
  b. Publication year
  c. Country of origin
  d. Funding information
  e. Purpose/primary aim(s) of study
2. Study design
3. Study population characteristics
  a. Sample size
  b. Age
  c. Sex
  d. Race/Ethnicity
  e. Presence of comorbidities
  f. Weight
  g. BMI
4. Prevention strategy characteristics
  a. Type of strategy (e.g., intervention, policy…)
  b. Locale of strategy: individual, single-site (e.g., clinic, institution), multi-site, local (e.g., town, city), regional, national
  c. Description of strategy
  d. Implementation duration
  e. Start date
  f. End date
  g. Comparator: none, same population, control site or location, control not specified
  h. Description of comparator
5. Outcomes evaluated
  a. Duration of follow-up for outcomes
  b. Start date of outcome follow-up
  c. End date
  d. Outcome ascertainment description
  e. BMI
  f. Body weight
  g. Waist Circumference/Waist-to-Hip Ratio
  h. Prevalence of overweight/obesity
  i. Cardiometabolic risk factors (e.g., lipids, blood pressure, fasting glucose)
    - Pre-post estimates, time trends
    - Single timepoint
    - Change
  j. Other outcomes of interest
    - Program direct costs
    - Indirect/societal costs
    - Cost-effectiveness (Quality-adjusted life years, disability-adjusted life years, incremental cost-effectiveness ratio)
    - Adherence
    - Implementation metrics
    - Qualitative
    - Others, please specify
6. Results
  a. Effectiveness of these strategies as described including population-level summary statistics and/or indicators of obesity and/or weight gain at the individual-level, when appropriate.

## Evidence Synthesis

We will generate a table(s) providing the key descriptive information of all included articles. Separately, we will present country-specific obesity prevention strategies and summarize the available evidence for their effectiveness. We will also provide outcome data by similar strategy types across countries. We will qualitatively describe strategies that have demonstrated success within and between the countries of interest.

## Discussion

This scoping review will provide a comprehensive summary of available evidence for interventions, policies, or dietary or other lifestyle guidelines that have been implemented in Italy, France, and Japan. The strategies with evidence to support their effectiveness will inform potential prevention efforts for consideration in the US. A critical next step will be to identify barriers to adopting them in the US. Our review will also examine the extent to which the successful international prevention efforts have already been attempted in the US, and offer potential reasons for what might explain their lack of success in the US context.

## Search term

### PUBMED

<u>Search Dates</u>: January 1st, 1980 - September 25th, 2023

<u>Search Strategy 1</u>:

(“Energy Intake”[Mesh] OR “Diet, Healthy”[Mesh] OR “Portion Size”[Mesh] OR “Exercise”[Mesh] OR caloric[tiab] OR calories[tiab] OR healthy diet*[tiab] OR portion size*[tiab] OR exercise[tiab] OR physical activity[tiab] OR walk[tiab] OR walking[tiab])

AND

(“Body Weight”[Mesh:NoExp] OR “Weight Loss”[Mesh:NoExp] OR “Body Mass Index”[Mesh] OR body weight[tiab] OR weight loss[tiab] OR body mass index[tiab])

AND

(“Italy”[Mesh] OR “France”[Mesh] OR “Japan”[Mesh] OR Italian*[tiab] OR France[tiab] OR French[tiab] OR Japan[tiab] OR Japanese[tiab])

<u>Search Strategy 2:</u>

(“Overweight/prevention and control”[Mesh] OR prevent obesity[tiab] OR obesity prevention[tiab] OR preventing obesity[tiab] OR prevention of obesity[tiab]) AND (“Italy”[Mesh] OR “France”[Mesh] OR “Japan”[Mesh] OR Italy[tiab] OR Italian*[tiab] OR France[tiab] OR French[tiab] OR Japan[tiab] OR Japanese[tiab])

### EMBASE

<u>Search Dates</u>: January 1st, 1980 - September 25th, 2023

<u>Search Strategy</u>: ‘obesity prevention’ AND (‘italy’/exp OR ‘italy’ OR ‘france’/exp OR ‘france’ OR ‘japan’/exp OR ‘japan’)

### COCHRANE

<u>Search Dates:</u>January 1st, 1980 - September 25th, 2023

<u>Search Strategy</u>:

#1 MeSH descriptor: [Energy Intake] explode all tree

#2 MeSH descriptor: [Diet, Healthy] explode all trees

#3 MeSH descriptor: [Portion Size] explode all trees

#4 MeSH descriptor: [Exercise] explode all trees

#5 MeSH descriptor: [Energy Intake] explode all trees

#6 MeSH descriptor: [Body Weight] explode all trees

#7 MeSH descriptor: [Weight Loss] explode all trees

#8 MeSH descriptor: [Body Mass Index] explode all trees

#9 MeSH descriptor: [Italy] this term only

#10 MeSH descriptor: [France] this term only

#11 MeSH descriptor: [Japan] this term only

#12 (#1 OR #2 OR #3 OR #4 OR #5) AND (#6 OR #7 OR #8) AND (#9 OR #10 OR #11)

## Data Availability

All data produced in the present work are contained in the manuscript.

## References

1. CDC. Adult Obesity Facts. Accessed September 2, 2023, https://www.cdc.gov/obesity/data/adult.html

2. Bray GA. Medical consequences of obesity. The Journal of clinical endocrinology & metabolism. 2004;89(6):2583–2589.

3. Luppino FS, de Wit LM, Bouvy PF, et al. Overweight, obesity, and depression: a systematic review and meta-analysis of longitudinal studies. Archives of general psychiatry. 2010;67(3):220–229.

4. Sarwer DB, Polonsky HM. The psychosocial burden of obesity. Endocrinology and Metabolism Clinics. 2016;45(3):677–688.

5. Cawley J, Biener A, Meyerhoefer C, et al. Direct medical costs of obesity in the United States and the most populous states. Journal of managed care & specialty pharmacy. 2021;27(3):354–366.

6. Gallus S, Lugo A, Murisic B, Bosetti C, Boffetta P, La Vecchia C. Overweight and obesity in 16 European countries. European journal of nutrition. 2015;54:679–689.

7. Yoshiike N, Miyoshi M. Epidemiological aspects of overweight and obesity in Japan--international comparisons. Nihon Rinsho Japanese Journal of Clinical Medicine. 2013;71(2):207–216.

8. Mahood Q, Van Eerd D, Irvin E. Searching for grey literature for systematic reviews: challenges and benefits. Research synthesis methods. 2014;5(3):221–234.

